# The final blink: intact eyeblink conditioning in isolated dystonia

**DOI:** 10.1101/2021.07.26.21260809

**Authors:** Anna Sadnicka, Lorenzo Rocchi, Anna Latorre, Elena Antelmi, James Teo, Isabel Pareés, Britt S Hoffland, Kristian Brock, Katja Kornysheva, Mark J Edwards, Kailash P Bhatia, John C Rothwell

## Abstract

**Background:** Impaired eyeblink conditioning is often cited as evidence for cerebellar dysfunction in isolated dystonia yet the results from individual studies are conflicting and underpowered.

**Objective:** To systematically examine the influence of dystonia, dystonia subtype and clinical features over eyeblink conditioning within a statistical model which controlled for the covariates age and sex.

**Methods:** Original neurophysiological data from all published studies were shared to an age and sex matched control group. Two raters blinded to participant identity scored all recordings (6732 trials). After higher inter-rater agreement was confirmed, mean conditioning per block was entered into a mixed repetitive measures model.

**Results:** Much variability of eyeblink conditioning was seen in both groups. All dystonia versus controls showed no difference in conditioning (p=0.517). Analysis of dystonia subgroup, with age and sex as covariates did not reveal evidence that eyeblink conditioning is impaired in specific subtypes (cervical dystonia, DYT-*TOR1A*, DYT-*THAP1* or focal hand dystonia). The presence of tremor did not significantly influence levels of eyeblink conditioning.

**Conclusions:** Collaborative efforts such as this article allow larger number of patients to be evaluated and provide more balanced estimates of disease effects over experimental outcomes. Intact eyeblink conditioning is against a global cerebellar learning deficit in isolated dystonia. Precise mechanisms for how the cerebellum interplays mechanistically with other key neuroanatomical nodes within the dystonic network remains an open research question.

## Introduction

Dystonia is a hyperkinetic movement disorder characterized by involuntary sustained muscle contractions which lead to twisting and repetitive movements or abnormal postures^1^. Traditionally, considered a disorder of basal ganglia function, recent research has pointed to abnormalities of multiple brain regions and within this wider sensorimotor network the cerebellum is thought to be a key node^2-4^.

Eyeblink conditioning is a cerebellar dependent experimental paradigm that has been traditionally used to study cerebellar function. Pavlovian by design, a biologically potent stimulus (the unconditioned stimulus) is paired with a previously neutral stimulus (the conditioning stimulus). Experimentally in humans the unconditioned stimulus is typically supraorbital nerve stimulation causing a blink (the unconditioned response). The conditioning stimulus (usually an auditory tone) always occurs before the unconditioned stimulus and with time the tone alone yields a conditioned blink (Fig 1a). Informatively, eyeblink conditioning can be adapted and tested across species and elements of the paradigm can be specifically mapped to the function of individual cells within the cerebellar circuitry^5^. For example, the magnitude of conditioned eyeblink responses correlates on a trial-by-trial manner to the level of firing of Purkinje cells (magnitude of simple spike suppression)^5,6^. Eyeblink conditioning is therefore an attractive experimental method with the potential to map behavioral outcomes to the cerebellar micro-circuitry.

**Figure 1.**
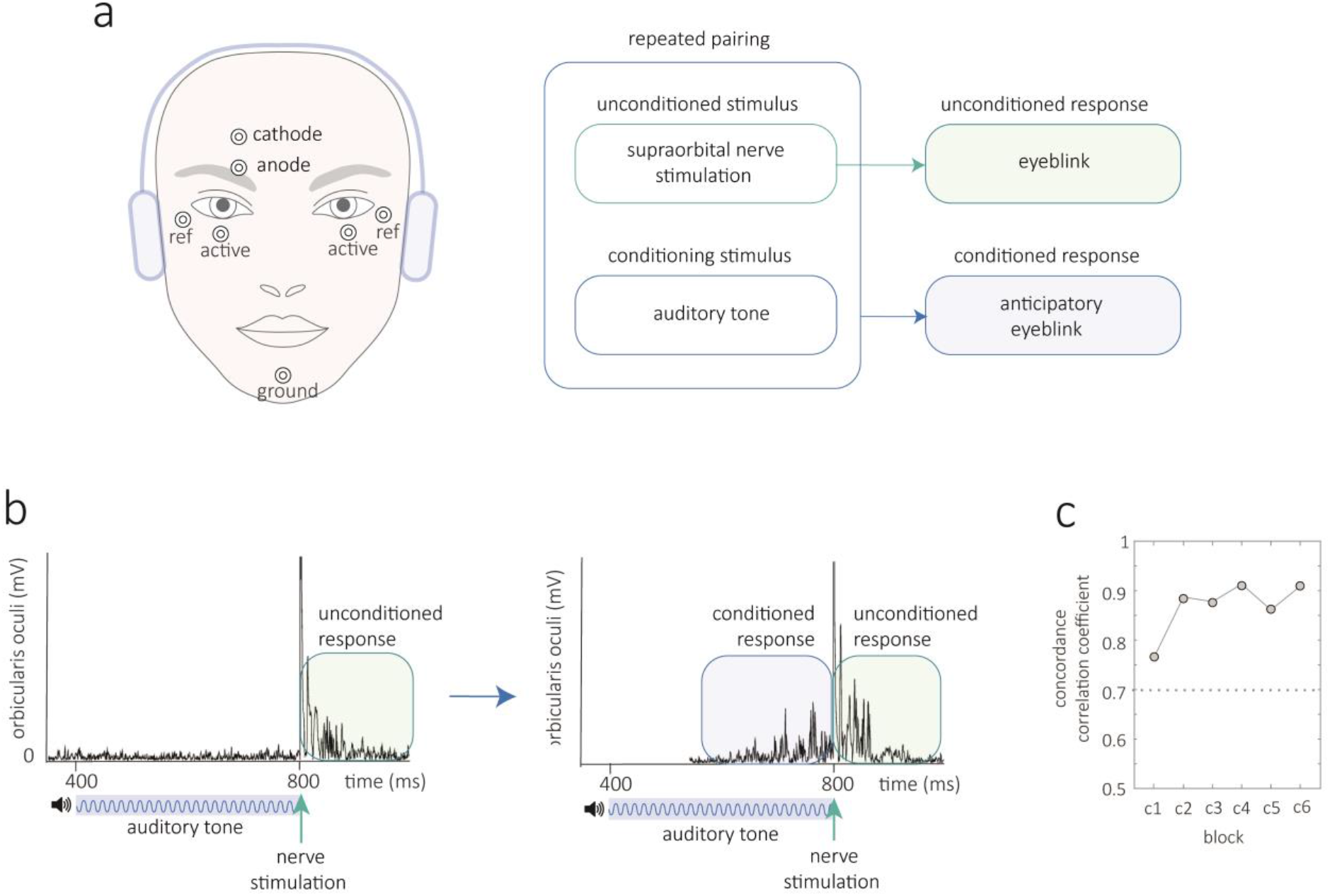
Eyeblink conditioning in humans. Method a| The unconditioned stimulus consists of electrical stimulation to the supraorbital nerve at 800ms which causes a blink, the unconditioned response. The conditioning stimulus is an auditory tone that starts at 400ms with a duration of 400ms. With repeated pairings a conditioned blink response emerges prior to supraorbital nerve stimulation. b| Rectified electromyographic traces from a single trial early with no conditioned response and a later trial when conditioning has developed. c| Two assessors scored the number of conditioned responses in every trial whilst blinded to the participants’ identity. Their post hoc concordance correlation coefficient was excellent across all blocks (values >0.7 are considered reasonable, see supplementary methods for detail).

Impaired eyeblink conditioning is widely cited as evidence of functional cerebellar disturbance in the dystonia literature, yet, collectively findings across studies are conflicting. A preliminary study examined a mixed group of cervical dystonia and focal hand dystonia and found that both had lower levels of eyeblink conditioning^7,8^. However, a later study documented normal eyeblink conditioning in cervical dystonia and reduced conditioning was only seen if there was coexisting head tremor^8^. Eyeblink conditioning in genetic subtypes appear to be either normal in DYT-*TOR1A* related dystonia or high in DYT-*THAP1* related dystonia (unless an age matched control group is used)^9^. Thus low, normal and high levels of eyeblink conditioning have been reported across subtypes of isolated dystonia. There are many potential reasons for the differences observed across studies. Firstly, levels of eyeblink conditioning in the healthy population are highly variable. Clinical studies are therefore vulnerable to inconsistency if underpowered and uncontrolled covariates such as sex and age can confound^10,11^. Recent research also points to the subtypes of isolated dystonia having unique etiologies and/or neuroanatomical substrates and thus uniform patterns of eyeblink conditioning behavior across the subtypes are not necessarily anticipated^12^.

This current study capitalized on an unusual opportunity in isolated dystonia to collate raw data from individual studies. Our motivation was to examine whether isolated dystonia as a group is associated with altered eyeblink conditioning. We also evaluated whether there is any evidence that eyeblink conditioning is altered across subtypes of isolated dystonia and whether tremor or other clinical features influence conditioning. Clinical questions were probed within a statistical model that included age and sex as covariates, as young age and female sex are associated with higher levels of conditioning.

## Method

This collaborative project identified all studies that have examined eyeblink conditioning in isolated dystonia. Original neurophysiological data were shared and a sex and age matched control group were collected. Two raters blinded to participant identity scored all recordings (controls n=50, dystonia n=52, trials=6732). After high inter-rater agreement was confirmed (Fig 1c), mean conditioning per block was entered into a mixed repetitive measures model to evaluate the influence of sex, age, dystonia, dystonia subtype and clinical features. Full details of the methods are given in the supplementary information.

## Results

A wide range of conditioning behavior was observed in both controls and patients with dystonia (Fig 2a). Some subjects did not exhibit any conditioned responses (0% conditioning across all blocks) whereas others had conditioned responses counted in early blocks. The two groups had very similar demographic features in terms of age (t(99)=0.0901, *p*=.928) and sex (equivalent proportions in each group). A plot of mean rates of conditioning by group (control, all dystonia) showed that eyeblink conditioning was similar to controls if all types of dystonia were considered as a group (Fig 2b, *p*=.517). Younger age was associated with higher conditioning (Fig 2b, *p*=.031). Higher levels of conditioning were also seen with female sex; a finding that was not significant in this study (*p*=.143), but has been observed reliably in previous studies^11^.

**Figure 2.**
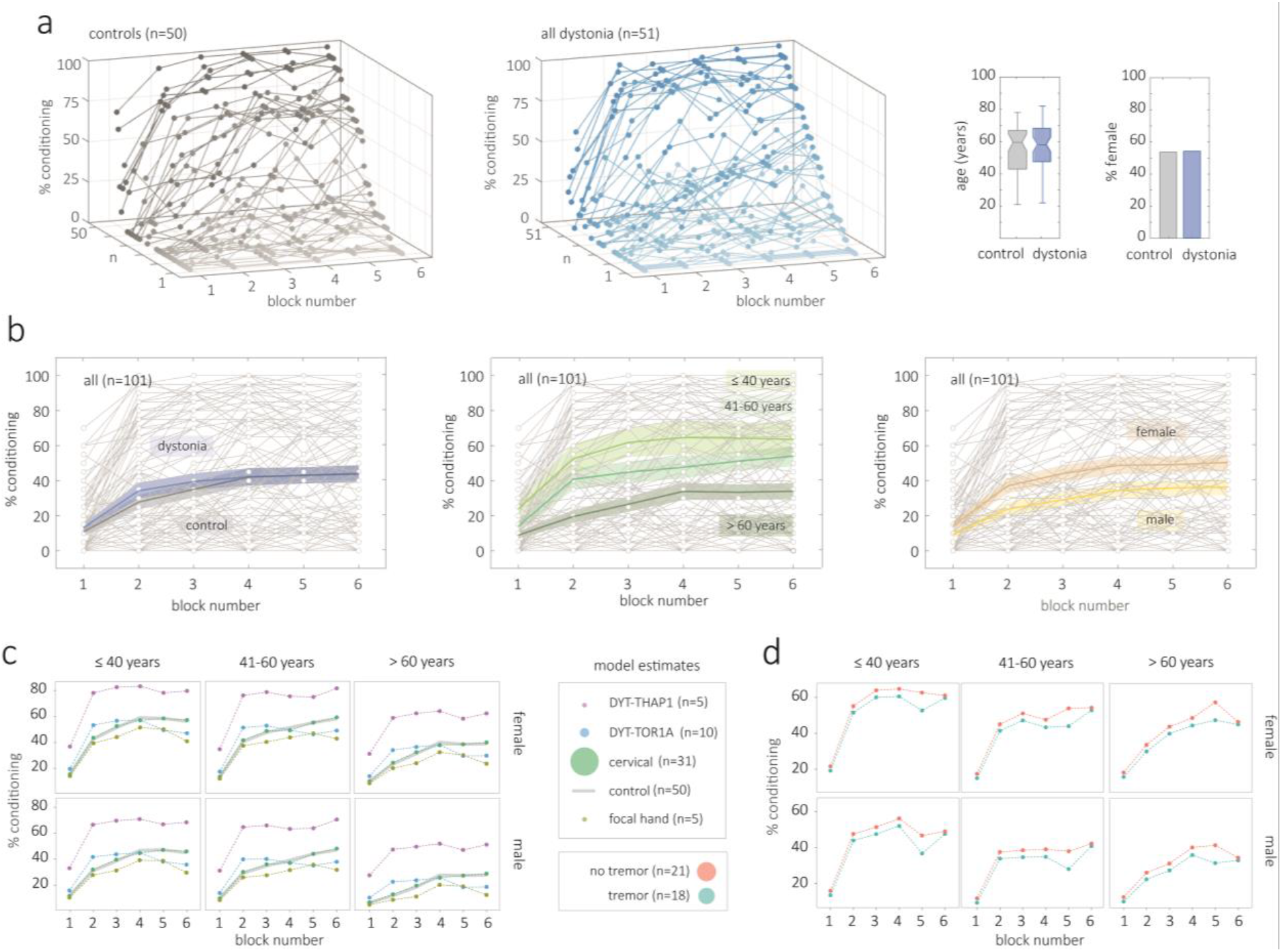
a| The range of conditioning across individuals is demonstrated in three dimensional plots for controls (n=50) and dystonia (n=51). Block number 1-6 is shown on the x-axis, % conditioning on the vertical y-axis, and participant number on the z-axis. For each group the profile of responses is sorted according to the % conditioning achieved by the final block. % conditioning for each block is marked by a filled circle and connected by a thin line of the same color. The groups were matched for age and sex. b| For each plot, all participants (n=101) are plotted in pale gray in the background to illustrate degree of variability. Each graph then shows the group mean and shaded standard error when grouped by presence of dystonia, age range and sex (control=grey, dystonia=blue, <=40 years = light green, 41-60 years = green, >60 years = dark green, female=tan, male=yellow). c and d | On the y-axis of each plot the model counterfactual is plotted, reflecting the model’s best estimate for conditioning behavior by block for each age (horizontal panels) and sex category (vertical panels). The shared key of each line plots shows the marker size proportional to the number of participants. In c| controls are plotted with a partially transparent grey line to aid visualization of overlapping lines.

Model estimates for conditioning behavior for dystonia subtype across different age and sex categories are plotted in Fig 2c. The mean difference between patients with DYT-THAP1 dystonia and controls was 26 units but there were only five patients in this in this subgroup (1 unit = 1 % conditioning, *p*=.049). Mean differences between dystonia subgroup and controls were < 10 units (all *p*-values >.5).

Finally, we reviewed key clinical parameters to assess whether they influenced levels of conditioning. We examined disease features (severity, duration, tremor) and active treatments at the time of study (botulinum toxin injections and/or the medications trihexyphenidyl and clonazepam). The presence of tremor did not affect eyeblink conditioning in the ‘all’ dystonia group (Figure 2d, *p*=.943) or the cervical dystonia subgroup (*p*=.514). No other disease features or treatments at the time of study exhibited any clear effect over conditioning (all *p*-values > 0.5). Based on the variance we observed in the final conditioning block we performed sample size calculations as a guide for future studies. The standard deviation in controls was 35.3%, standard deviation in dystonia 32.7%, with a mean standard deviation of 34.0%. Therefore, in order to detect a difference in conditioning of 5% a sample size of 739 per group is required. A difference of conditioning of 10% requires 185 per group, 20% requires 46 per group and 30% requires 21 per group.

## Discussion

Our results do not find evidence that isolated dystonia and the subtypes of isolated dystonia are associated with changes in eyeblink conditioning relative to controls. Our data suggest a revision to the idea that lower levels of eyeblink conditioning are a feature of focal dystonia^7^. As eyeblink conditioning is often discussed as a proxy for cerebellar function, our findings have implications for how we define cerebellar involvement in dystonia pathophysiology.

Multiple convergent lines of evidence point suggest that the cerebellum is involved in the pathophysiology of dystonia. For example, in humans cerebellar lesions can cause symptomatic/acquired dystonia, in genetic animal models cerebellar perturbations appear sufficient to generate dystonia and in focal dystonia imaging studies consistently point to cerebellar abnormalities^13-15^. Therefore, there is collective evidence that the cerebellum is a key node within dystonic networks and cerebellar dysfunction may play a dominant role is some subtypes. However, we are still far from identifying specific cerebellar mechanisms. How do our results contribute to this discussion?

Firstly, in general terms, our results suggest that there is a relative subtlety to any cerebellar dysfunction in genetic/idiopathic isolated dystonia. Neurodegenerative disorders such as spinocerebellar ataxia (type 6) affect the cerebellum relatively uniformly and are associated with broad functional impairments ranging from delayed eyeblink conditioning and force adaptation in reaching^16-20^. More circumscribed cerebellar deficits, e.g. caused by cerebellar stroke, can produce highly task-specific impairments, such as abnormal adaptation to force but not to visuomotor adaptation and vice versa^21,22^. Forcefield and visuomotor adaptation paradigms have also been studied in isolated dystonia subtypes are there is not strong evidence that this movement calibration/computation is impaired^23-26^. Therefore, the absence of clear deficits in dystonia to these archetypal cerebellar paradigms and absence of overt cerebellar signs does suggest that there is a subtlety to cerebellar involvement.

To more precisely interpret our results, it is useful return to the observation that different elements of eyeblink conditioning have been mapped to the firing of individual cells within the cerebellar circuitry. As the cerebellar micro-circuitry has a highly homogenous nature this has given rise to the idea that the cerebellum performs the same function at the algorithmic level across diverse domains (‘universal transform’)^27^. In such a scenario the broad functional heterogeneity of different cerebellar regions across motor control, perception, language, and cognition is primarily determined by connectivity patterns to cortical and subcortical targets rather than the micro-circuitry^27^. Therefore, if we interpret our results in line with the ‘universal cerebellar transform’ theory, most simply, confirmation of the integrity of the general micro-circuitry and plasticity of the cerebellum via eye-blink conditioning, could be seen as a sample of global cerebellar health across all functional domains. However, there are obvious caveats to the assumption eyeblink conditioning performance reflects cerebellar health and cerebellar processing in its entirety. For example, an alternate viewpoint is that the complete range of cerebellar function entails multiple specialized algorithms across different cerebellar regions^27^. In this scenario the same underlying circuit implements functionally distinct algorithms sub serving different functional modules and tasks (‘multiple functionality’)^16,27^. Correspondingly, it may be that eyeblink conditioning is not tapping into the specific functionality at play in dystonia pathophysiology, a specific algorithm that underwrites the dystonic phenotype. Overall, mechanistically, it remains largely unknown how the cerebellum interplays with other implicated neuroanatomical nodes in the pathophysiology of isolated dystonia. In the future, we are likely to lean on techniques that evaluate multiple nodes within the network simultaneously in order to gain broader, less one-dimensional insight. Synergistic and/or compensatory interactions between nodes may define novel mechanisms in dystonia.

We also did not find evidence to support the hypothesis that tremor and other clinical features such as disease severity or medications influence conditioning. Statistically, when controlling for the covariates age and sex we did not replicate findings in a previous study in cervical dystonia in which impaired eyeblink conditioning was linked to those with head tremor^8^. Similar to dystonia, a network model of the pathophysiology of tremor is proposed in which the olivo-cerebellar system is thought to be critical. Patients with essential tremor have had both low and normal levels of eyeblink conditioning documented depending on the paradigm used. However, as cerebellar cells show similar responses across paradigms, further studies are needed to establish the fidelity of findings^28^. Of note, low levels of cerebellar conditioning do not appear to be a marker of any tremor syndrome as eyeblink conditioning appears to be normal in tremor associated with neuropathy^29^.

Limitations of our study are that the numbers within subgroups of dystonia were small. Although we extended previous conclusions by modeling the covariates age and sex and making comparisons to the larger control group, we were underpowered to reliably evaluate subgroups. Indeed, our sample size calculations reveal large studies are required to confidently assess for changes in eyeblink conditioning behavior and most individual studies in the literature are not adequately powered. For example, to detect a 20 % difference in the level of conditioning across groups a minimum of 46 participants per group is required. This does bring into question whether eyeblink conditioning is useful to study in heterogeneous disease groups given its inherent variability. Also, in the context of our study, although identical methods and equipment are documented and confirmed amongst authors, some of the reasons for variability of conditioning response are still likely to reflect non-biological influences such as differences in experimental technique across investigators

In summary, eyeblink conditioning in isolated dystonia appears intact and clinical features of dystonia such as tremor did not significantly modulate the level of conditioning observed. Collaborative efforts such as this article allow larger number of patients to be evaluated to provide more balanced estimates of disease effects over experimental paradigms. Normal eyeblink conditioning is against a global cerebellar learning deficit in isolated dystonia. Future studies are required to elucidate exactly how cerebellar involvement interplays with other key neuroanatomical nodes implicated in the dystonic network.

## Data Availability

NA

## Acknowledgements

The authors thank Cambridge Electronic Design for providing remote licenses to investigators which allowed analysis to continue whilst labs were closed during the pandemic.

## Authors’ Roles

AS 1ABC 2ABC 3AB

LR 1ABC 2A 3B

AL 1ABC 3B

EA 1C 3B

JT 1C 3B

BH 1C 3B

IP 1C 3B

KB 2A 2B 3B

KK 1B 3B

ME 1A 3B

JR 1A 3B

KB 1A 3B

## Financial Disclosures of all authors (for preceding 12 months)

Sadnicka: Chadburn Clinical Lectureship in Medicine. Kornysheva: Academy of Medical Sciences Springboard Award (SBF006\1052)

## References

1 Albanese, A. et al. Phenomenology and classification of dystonia: a consensus update. Mov Disord 28, 863–873, doi:10.1002/mds.25475 (2013).

2 Berardelli, A. et al. The pathophysiology of primary dystonia. Brain 121 (Pt 7), 1195–1212, doi:10.1093/brain/121.7.1195 (1998).

3 Marsden, C. D., Obeso, J. A., Zarranz, J. J. & Lang, A. E. The anatomical basis of symptomatic hemidystonia. Brain 108 (Pt 2), 463–483 (1985).

4 Neychev, V. K., Gross, R. E., Lehericy, S., Hess, E. J. & Jinnah, H. A. The functional neuroanatomy of dystonia. Neurobiol Dis 42, 185–201, doi:10.1016/j.nbd.2011.01.026 (2011).

5 ten Brinke, M. M. et al. Evolving Models of Pavlovian Conditioning: Cerebellar Cortical Dynamics in Awake Behaving Mice. Cell Rep 13, 1977–1988, doi:10.1016/j.celrep.2015.10.057 (2015).

6 Ten Brinke, M. M., Boele, H. J. & De Zeeuw, C. I. Conditioned climbing fiber responses in cerebellar cortex and nuclei. Neurosci Lett 688, 26–36, doi:10.1016/j.neulet.2018.04.035 (2019).

7 Teo, J. T., van de Warrenburg, B. P., Schneider, S. A., Rothwell, J. C. & Bhatia, K. P. Neurophysiological evidence for cerebellar dysfunction in primary focal dystonia. J Neurol Neurosurg Psychiatry 80, 80–83, doi:10.1136/jnnp.2008.144626 (2009).

8 Antelmi, E. et al. Impaired eye blink classical conditioning distinguishes dystonic patients with and without tremor. Parkinsonism Relat Disord 31, 23–27, doi:10.1016/j.parkreldis.2016.06.011 (2016).

9 Sadnicka, A. et al. All in the blink of an eye: new insight into cerebellar and brainstem function in DYT1 and DYT6 dystonia. Eur J Neurol 22, 762–767, doi:10.1111/ene.12521 (2015).

10 Solomon, P. R., Pomerleau, D., Bennett, L., James, J. & Morse, D. L. Acquisition of the classically conditioned eyeblink response in humans over the life span. Psychol Aging 4, 34–41, doi:10.1037//0882-7974.4.1.34 (1989).

11 Lowgren, K. et al. Performance in eyeblink conditioning is age and sex dependent. PLoS One 12, e0177849, doi:10.1371/journal.pone.0177849 (2017).

12 Lungu, C. et al. Defining research priorities in dystonia. Neurology 94, 526–537, doi:10.1212/WNL.0000000000009140 (2020).

13 Jinnah, H. A., Neychev, V. & Hess, E. J. The Anatomical Basis for Dystonia: The Motor Network Model. Tremor Other Hyperkinet Mov (N Y) 7, 506, doi:10.7916/D8V69X3S (2017).

14 Prudente, C. N., Hess, E. J. & Jinnah, H. A. Dystonia as a network disorder: what is the role of the cerebellum? Neuroscience 260, 23–35, doi:10.1016/j.neuroscience.2013.11.062 (2014).

15 Fremont, R., Tewari, A., Angueyra, C. & Khodakhah, K. A role for cerebellum in the hereditary dystonia DYT1. Elife 6, doi:10.7554/eLife.22775 (2017).

16 Timmann, D., Gerwig, M., Frings, M., Maschke, M. & Kolb, F. P. Eyeblink conditioning in patients with hereditary ataxia: a one-year follow-up study. Exp Brain Res 162, 332–345, doi:10.1007/s00221-004-2181-x (2005).

17 Maschke, M., Gomez, C. M., Ebner, T. J. & Konczak, J. Hereditary cerebellar ataxia progressively impairs force adaptation during goal-directed arm movements. J Neurophysiol 91, 230–238, doi:10.1152/jn.00557.2003 (2004).

18 Gerwig, M. et al. Comparison of eyeblink conditioning in patients with superior and posterior inferior cerebellar lesions. Brain 126, 71–94, doi:10.1093/brain/awg011 (2003).

19 Gerwig, M. et al. Amplitude changes of unconditioned eyeblink responses in patients with cerebellar lesions. Exp Brain Res 155, 341–351, doi:10.1007/s00221-003-1731-y (2004).

20 Gerwig, M. et al. Timing of conditioned eyeblink responses is impaired in cerebellar patients. J Neurosci 25, 3919–3931, doi:10.1523/JNEUROSCI.0266-05.2005 (2005).

21 Donchin, O. et al. Cerebellar regions involved in adaptation to force field and visuomotor perturbation. J Neurophysiol 107, 134–147, doi:10.1152/jn.00007.2011 (2012).

22 Burciu, R. G. et al. Structural correlates of motor adaptation deficits in patients with acute focal lesions of the cerebellum. Exp Brain Res 232, 2847–2857, doi:10.1007/s00221-014-3956-3 (2014).

23 Sadnicka, A. et al. Normal motor adaptation in cervical dystonia: a fundamental cerebellar computation is intact. Cerebellum 13, 558–567, doi:10.1007/s12311-014-0569-0 (2014).

24 Lange, L. M. et al. Genotype-Phenotype Relations for Isolated Dystonia Genes: MDSGene Systematic Review. Mov Disord 36, 1086–1103, doi:10.1002/mds.28485 (2021).

25 Katschnig-Winter, P. et al. Motor sequence learning and motor adaptation in primary cervical dystonia. J Clin Neurosci 21, 934–938, doi:10.1016/j.jocn.2013.08.019 (2014).

26 Sadnicka, A. et al. High motor variability in DYT1 dystonia is associated with impaired visuomotor adaptation. Sci Rep 8, 3653, doi:10.1038/s41598-018-21545-0 (2018).

27 Diedrichsen, J., King, M., Hernandez-Castillo, C., Sereno, M. & Ivry, R. B. Universal Transform or Multiple Functionality? Understanding the Contribution of the Human Cerebellum across Task Domains. Neuron 102, 918–928, doi:10.1016/j.neuron.2019.04.021 (2019).

28 Solbach, K., Oostdam, S. J., Kronenbuerger, M., Timmann, D. & Gerwig, M. Long Trace Eyeblink Conditioning Is Largely Preserved in Essential Tremor. Cerebellum 18, 67–75, doi:10.1007/s12311-018-0956-z (2019).

29 Saifee, T. A. et al. Tremor in Charcot-Marie-Tooth disease: No evidence of cerebellar dysfunction. Clin Neurophysiol 126, 1817–1824, doi:10.1016/j.clinph.2014.12.023 (2015).

