## Supplementary methods for "The final blink: intact eyeblink conditioning in isolated dystonia"

PubMed and MEDLINE were searched with the terms dystonia AND ((eyeblink conditioning) OR (associative learning)) and identified three studies for inclusion (Supplementary Fig 1). All followed the same experimental paradigm and had been tested within labs at University College London between 2009-2019. Each study had a different lead investigator and new data collected in each paper came from a discrete study. Across studies, patients receiving botulinum toxin injections had been tested at the end of the therapeutic window (at least 12 weeks after last injection). Demographic and neurophysiological data files were collated. For each participant demographic age and sex were noted. For the dystonia group, subtype of dystonia, duration of symptoms, severity of dystonia, presence of tremor, medication and botulinum toxin use was also recorded (supplementary Table 1). As many of these studies had been compared to a historical, small control group (n=8) we collected further control data. This study had been approved by the local ethics committee and written informed consent was obtained.

**Supplementary Figure 1. Study and participant identification** A: Eight studies that have examined eyeblink conditioning and dystonia were identified. Studies were excluded if they did not examine isolated dystonia (three studies): myoclonus dystonia or fixed/functional dystonia[5-7]. Another paper did not contain novel data in isolated dystonia[8]. Neurophysiological data for a final study were not available and this would have contributed eight further patients with cervical dystonia, same historical control group[9]. B: the number of patients and controls for each study are noted[1-3]. Data for a single patient with dystonia were not available from the Antelmi study (n=24). Further new control data were collected for this study.

a

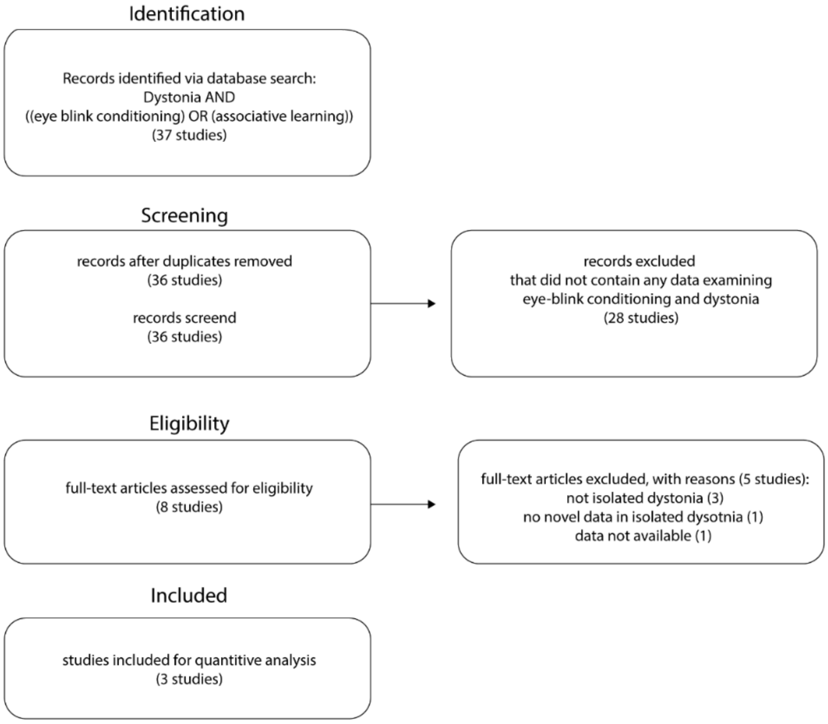


b

| Study | Dystonia group | Control group |
|---|---|---|
| Teo | focal hand (n=5)<br>cervical (n=7) | novel (n=8) |
| Antelmi | cervical (n=24) | historical (n=8) |
| Sadnicka | DYT-TOR1A (n=11)<br>DYT-THAP1 (n=5) | historical (n=8)<br>novel (n=6) |
| Current study | no novel | novel (n=36) |
| Total | all dystonia (n=52) | all control (n=50) |

The eyeblink conditioning paradigm was the same across studies (Fig 1a)[1-3]. Eye blinks were captured by surface electromyographic (EMG) electrodes over right and left orbicularis oculi muscles. Signal was amplified (gain 2000), bandpass filtered (20Hz–3000Hz) and digitized (5kHz). The unconditioned stimulus (US) was an electrical stimulus (200µsec, 5 times sensory threshold) to the supraorbital nerve at the termination of the conditioning stimulus which elicited a blink reflex (UR). Supraorbital nerve was applied with chloride disc surface electrodes (electrodes located at the right supraorbital foramen and 2 cm above). The conditioning stimulus (CS) was a loud (~70dB), 2000Hz, 400ms tone played via binaural headphones. Repeated pairs of conditioning and unconditioned stimuli yielded conditioned blink responses (CR) occurring before supraorbital nerve stimulation (Fig 1b). There were six 'conditioning' blocks of eleven trials with the following structure: 9 x CS-US, 1 x US and 1 x CS. The CS-only confirmed that CRs were acquired independent of US. EMG bursts were regarded as CRs if latency was >200ms after onset of CS but before the US. We have not reported data on the seventh block which measured extinction with eleven CS-only trials as this was not our primary research question.

Data for 50 controls and 52 patients were scored. Data were analyzed offline analysis using Cambridge Electronic Design signal software (CED). Every trial (11 trials x 6 blocks x 102 participants = 6732 trials) was assessed for the presence of a conditioned response by two assessors blinded to the participants identity (authors AS and LR). One participant with dystonia was excluded due to irregularities with the experimental paradigm. In order to assess the agreement across assessors the concordance correlation coefficient was calculated for each block of the experiment (Fig 2c)

$$\text{concordance correlation coefficent} = \frac{2\,\sigma_{12}}{\sigma_1^2 + \sigma_2^2 + (\mu_1 + \mu_2)^2}$$

where $\sigma_{12}$ is the covariance between the two distributions and $\sigma_x^2$, $\mu_x$ are the variance and mean of distribution x respectively. This quantifies the similarity between distributions based on their covariance and average values; concordance correlation coefficient values >0.7 are considered reasonable[4]. High inter-rater agreement of scores across all blocks of conditioning assessed was found suggesting that levels of conditioning were reliably identified from data (Fig 1c). The mean % conditioning value for each block across the two assessors was subsequently used for data visualization and statistical analysis. In total data for 50 controls (27 females, mean age 55.2 years (SD 15.5 years)) and 51 individuals with dystonia (28 females, mean age 56.0 years (SD 15.1 years)) were entered into statistical analysis.

Supplementary Figure 2. a | A plot of the average score across the two assessors versus the different in scores between assessors for the final block (c6). It is possible to score between 0 (0% conditioning) and 10 (100% conditioning). The mean difference (shown by solid line) revealed an average difference of <1. b| In some participants, assessors had different thresholds for labelling conditioned responses as the 'difference between scorers' had a unilateral tail on this frequency plot (negative on this plot)

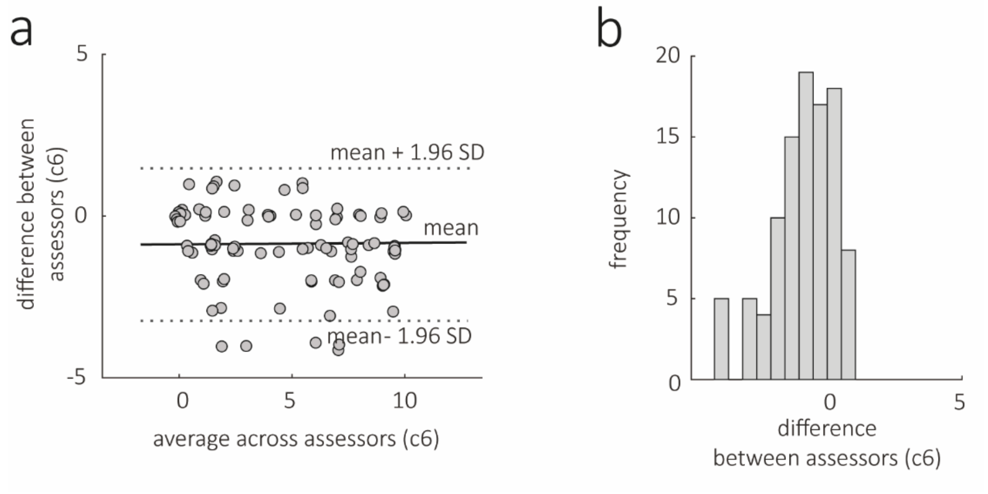

We used a mixed-effects model for repeated measures to analyze conditioning outcomes[10]. The model used variables to estimate the effects associated with age, sex, patient group, experimental block, and the intersections of age, sex and group with block. Treating the first block as baseline, exploratory data analysis revealed that post-baseline outcomes had approximately constant variance and serial correlation, irrespective of block number. This suggested that a compound symmetric covariance structure was appropriate and so patient-level intercepts were used in the model. Model checks revealed that the residuals were approximately independent, homoskedastic, and normally distributed, and that patient-level intercepts were approximately normal. Tests of difference between subgroups were made using likelihood ratio tests of nested models. Further fixed effects were added to this model to test the explanatory power of clinical variables. Tremor, botulinum toxin, trihexyphenidyl and clonazepam treatments were coded as either present or absent. Severity of dystonia was described as a percentage relative to the total of the scoring scale used for each subtype of dystonia. Duration of symptoms was estimated to the closest year.

Sample size estimates for the minimum size of each arm were made by extrapolating from the variability of conditioning levels observed in the final block 6:

$$\text{sample size} = \frac{2\sigma^2 \left(Z_{\frac{\alpha}{2}} + Z_{\beta}\right)^2}{d^2} = \frac{2\sigma^2\,(1.96 + 0.84)^2}{d^2} = \frac{16\,\sigma^2}{d^2}$$

where $\sigma$ is the standard deviation, d is the effect size and Z constants are calculated on the basis of desired significance level (type 1 error, alpha) and power (probability of rejecting null hypothesis when false). Constants were defined for a standard two sided comparison with a significance level of 5% and power of 80%[11]. All results are given to three significant figures.

### References


1 Teo, J. T., van de Warrenburg, B. P., Schneider, S. A., Rothwell, J. C. & Bhatia, K. P. Neurophysiological evidence for cerebellar dysfunction in primary focal dystonia. *J Neurol Neurosurg Psychiatry* **80**, 80-83, doi:10.1136/jnnp.2008.144626 (2009).

2 Antelmi, E. *et al.* Impaired eye blink classical conditioning distinguishes dystonic patients with and without tremor. *Parkinsonism Relat Disord* **31**, 23-27, doi:10.1016/j.parkreldis.2016.06.011 (2016).

3 Sadnicka, A. *et al.* All in the blink of an eye: new insight into cerebellar and brainstem function in DYT1 and DYT6 dystonia. *Eur J Neurol* **22**, 762-767, doi:10.1111/ene.12521 (2015).

4 Lin, L. I. A concordance correlation coefficient to evaluate reproducibility. *Biometrics* **45**, 255-268 (1989).

5 Popa, T. *et al.* The neurophysiological features of myoclonus-dystonia and differentiation from other dystonias. *JAMA Neurol* **71**, 612-619, doi:10.1001/jamaneurol.2014.99 (2014).

6 Weissbach, A. *et al.* Alcohol improves cerebellar learning deficit in myoclonus-dystonia: A clinical and electrophysiological investigation. *Ann Neurol* **82**, 543-553, doi:10.1002/ana.25035 (2017).

7 Janssen, S. *et al.* Normal eyeblink classical conditioning in patients with fixed dystonia. *Exp Brain Res* **232**, 1805-1809, doi:10.1007/s00221-014-3872-6 (2014).

8 Kojovic, M. *et al.* Secondary and primary dystonia: pathophysiological differences. *Brain* **136**, 2038-2049, doi:10.1093/brain/awt150 (2013).

9 Hoffland, B. S. *et al.* Cerebellum-dependent associative learning deficits in primary dystonia are normalized by rTMS and practice. *Eur J Neurosci* **38**, 2166-2171, doi:10.1111/ejn.12186 (2013).

10 Mallinckrod, C. H., Lane, P. W., Schnell, D., Peng, Y. & Mancuso, J. P. Recommendations for the Primary Analysis of Continous Endpoints in Longitudinal Clinical Trials. *Therapeutic Innovation & REgulatory Sceince* **42**, 16 (2008).

11 Campbell, M. J., Julious, S. A. & Altman, D. G. Estimating sample sizes for binary, ordered categorical, and continuous outcomes in two group comparisons. *BMJ* **311**, 1145-1148, doi:10.1136/bmj.311.7013.1145 (1995).
